# Infection patterns of endemic human coronaviruses in rural households in coastal Kenya

**DOI:** 10.1101/2020.11.03.20225441

**Authors:** Dickson Machira Nyaguthii, Grieven P. Otieno, Ivy K. Kombe, Dorothy Koech, Martin Mutunga, Graham F. Medley, D. James Nokes, Patrick K. Munywoki

**Affiliations:** Epidemiology and Demography Department, KEMRI-Wellcome Trust Research Programme, Kilifi, Kenya. P.O. Box 230-80108 Kilifi, Kenya; Department of Veterinary Public Health, Pharmacology and Toxicology, Faculty of Veterinary Medicine and Surgery, Egerton University, P.O. Box 536-20115, Egerton, Kenya; Centre for Mathematical Modelling of Infectious Disease, London School of Hygiene and Tropical Medicine, London, WC1H 9SH, United Kingdom; School of Life Sciences and Zeeman Institute for Systems Biology and Infectious Disease Epidemiology Research (SBIDER), University of Warwick, Coventry, CV4 7AL, United Kingdom

**Keywords:** Human coronavirus, OC43, NL63, 229E, Transmission, Households, Virus shedding

## Abstract

**Introduction:** The natural history and transmission patterns of endemic human coronaviruses are of increased interest following the emergence of severe acute respiratory syndrome coronavirus-2 (SARS-CoV-2).

**Methods:** In rural Kenya 483 individuals from 47 households were followed for six months (2009-10) with nasopharyngeal swabs collected twice weekly regardless of symptoms. A total of 16,918 swabs were tested for human coronavirus (hCoV) OC43, NL63 and 229E and other respiratory viruses using polymerase chain reaction.

**Results:** From 346 (71.6%) household members 629 hCoV infection episodes were defined with 36.3% being symptomatic: varying by hCoV type and decreasing with age. Symptomatic episodes (aHR=0.6 (95% CI:0.5-0.8) or those with elevated peak viral load (medium aHR=0.4 (0.3-0.6); high aHR=0.31 (0.2-0.4)) had longer viral shedding compared to their respective counterparts. Homologous reinfections were observed in 99 (19.9%) of 497 first infections. School-age children (55%) were the most common index cases with those having medium (aOR=5.3 (2.3 – 12.0)) or high (8.1 (2.9 - 22.5)) peak viral load most often generating secondary cases.

**Conclusion:** Household coronavirus infection was common, frequently asymptomatic and mostly introduced by school-age children. Secondary transmission was influenced by viral load of index cases. Homologous-type reinfection was common. These data may be insightful for SARS-CoV-2.

## Introduction

Four endemic species of human coronavirus (hCoV); HKU1, OC43, NL63 and 229E are widespread and associated primarily with mild acute respiratory illness [1]. Infections with endemic hCoVs are reportedly more severe in young children and the elderly [2,3]. In the last two decades, three new members of this virus family have emerged as human pathogens; severe acute respiratory syndrome coronavirus (SARS-CoV) [4], Middle East respiratory syndrome coronavirus (MERS-CoV) [5] and most recently SARS-CoV-2 [6]. The pandemic spread and continued circulation beyond the initial wave of infection suggests a potential for SARS-CoV-2 to become resident within the human population. A focus on the natural history and transmission characteristics of current little-studied endemic species of hCoV may give insight to the future behaviour of this emergent relative [7].

Using data from a study of 47 households in rural Kenya, we have previously reported baseline data on the occurrence of hCoV [8] and a detailed analysis of reinfection with hCoV-NL63 [9]. In the present study, we investigate the natural history of infection and transmission patterns of three endemic hCoV within these households.

## Materials and Methods

### The household data

This study utilizes data from a prospective household-based cohort study conducted in one administrative location within the Kilifi health and demographic surveillance system (KHDSS) [8,10] on the Kenyan coast. The study design and methods have been described elsewhere [8,11]. Briefly, with a primary objective of characterising ‘who infects whom’ with respiratory syncytial virus (RSV), households with an infant born after the end of the 2008/2009 RSV season (referred to as the study infant) and at least one elder sibling (aged <13 years) were enrolled. The study period spanned a complete RSV season from 8^th^ December 2009 to 5^th^ June 2010. Nasopharyngeal specimens (NPS) were collected from all household members irrespective of symptoms, once a week in the first four weeks and twice-a-week thereafter until the study end. A household was defined as members (who need not be related) of one or more building units who share the same cooking facility. The study had a good retention rate (>80%) of households and of individuals over the study period [11].

### Molecular testing of the NPS collections using multiplex RT-PCR assay

A previously described real time multiplex RT-PCR (mPCR) assay with targets for 15 respiratory viruses was used [12]. The target pathogens were human coronavirus (hCoV species (also called types) OC43, NL63 and 229E), RSV A and B, rhinovirus (RV), adenovirus (AdV), parainfluenza virus (types 1-4), influenza virus (types A, B and C) and human metapneumovirus (hMPV). A preliminary screen of the NPS showed the last three virus groups were uncommon during the surveillance period and hence not screened for the remainder of the NPS collections [8]. A specimen with a cycle threshold (Ct) value of ≤35.0 for a specific virus target was considered positive.

### Statistical analysis

Data analyses were undertaken in STATA Version 13.0 (StataCorp, College Station, Texas, USA). Descriptive statistics for continuous variables are presented as mean (± standard deviation) and median (interquartile range (IQR)). Categorical variables were summarised using counts and proportions and the chi-square test of association was used to examine the independence. The Mood’s median test was used to investigate equality of median times across levels of categorical variables. Two or more groups were compared using test for equality of proportions.

Type-specific individual hCoV infection episodes were defined as a period with positive mPCR result(s) of the same type with no more than 14 days apart [13]. Episodes where no samples were collected and tested for >7 days before or after the infection episode were considered left- or right-censored, respectively. An episode was considered symptomatic if the individual was identified with any of the following symptoms during the infection episode; cough, runny nose, sore throat, nasal flaring, indrawing, crackles, wheeze, fever, unable to feed, head nodding, lethargy, unable to talk, cyanosis or difficulty breathing. Co-infection was assigned when within the hCoV infection episode an NPS collection was mPCR positive for a different hCoV species or another of the viruses tested, namely; RSV, RV, or AdV. Detection of two or more individual infection episodes by the same hCoV type in a household within a span of 14 days constituted a household outbreak. For each household hCoV introduction, a primary (index) case was defined as the first person(s) to test positive for hCoV by mPCR while secondary case(s) were the rest of the members who are part of the same household outbreak. For individuals with multiple hCoV infection episodes, reinfections were classified as either homologous (same hCoV species) or heterologous (different hCoV species) with respect to previously detected species during the study period. As an example, if an individual has 3 infections in the temporal order OC43, NL63 and OC43, then the second infection episode would be heterologous to the first, and the third homologous to the first infection episode and heterologous to the second episode.

Durations of virus shedding were estimated using a midpoint method which was defined as the period starting midway between the first positive sample and the previous negative sample and ending midway between the last positive sample and the subsequent negative sample. Further details on this approach are provided elsewhere [13]. Kaplan Meir (KM) curves were used to describe the survival functions (time to end of virus shedding) by different categorical variables across the three endemic hCoV types. Adjusted hazard ratios (aHR) obtained from multivariable Cox proportional hazards (PH) models were used to estimate the influence of several factors on the duration of shedding and symptoms. Logistic regression models were used to identify risk factors for spread of infection from the primary cases to other household members. The risk factors considered were age, sex, household size, presence of respiratory symptoms, presence of other respiratory pathogens and peak viral load in an infection episode. The peak/highest viral load was defined as the lowest Ct value in an individual infection episode and was categorised into three levels; low (>=30), medium (20-29) and high (<20). To account for clustering either at individual or household level, robust cluster variance estimator was used in the Cox PH and logistic regression models discussed above.

## Results

### Baseline characteristics

A total of 483 individuals from 47 households had NPS collected over the six-month period. The mean number of household members was 10.5 (SD=6.5) classified into small (4-7 members), medium (8-16 members) and large (17-37 members). The median age of participants at the start of sampling was 10.7 years (IQR: 4.0 – 23.4). The cohort had 214 (44.3%) male participants. Of the 47 study infants, the average age at the start of the study was 3.9 (SD=2.6) months and 22 (46.8%) of the infants were males. A total of 16,918 NPS from 483 individuals were successfully tested for OC43, 229E and NL63. The median number of NPS collected from study participants was 41 (IQR: 30 - 44).

Of the 16918 samples tested, 1274 (7.5%) were positive for any of the three hCoV: 651 (3.8%), 418 (2.5%) and 241 (1.4%) samples were positive for OC43, NL63 and 229E, respectively. Seven (0.04%) NPS collections were positive for both OC43 and NL63, 17 (0.1%) were positive for both OC43 and 229E and 13 (0.08%) were positive for both NL63 and 229E. Only one sample was positive for all three hCoV tested. Higher individual crude attack rates (Table 1) were seen in school going children (129/169; 76.3%), males (160/214; 74.8%) and younger individuals (<1 year; 42/55, 76.4%, 1-4 years; 64/82, 78.0% and 5-14 years; 125/163, 76.7%) (Supplementary figure S1) and individuals residing small (75/95, 79.0%) and large households (97/124, 78.2%).

**Table 1:** Crude individual attack rates for the hCoV infections by various characteristics.

| Characteristics | Categories | Any hCoV |  |  | OC43 |  | NL63 |  | 229E |  |
| --- | --- | --- | --- | --- | --- | --- | --- | --- | --- | --- |
|  |  | *N* | *n* | % | *n* | % | *n* | % | *n* | % |
| **Overall** |  | 483 | 346 | 71.6 | 215 | 44.4 | 163 | 33.7 | 119 | 24.6 |
| **Age in years** | <1 | 55 | 42 | 76.4 | 30 | 54.6 | 16 | 29.1 | 16 | 29.1 |
|  | 1-4 | 82 | 64 | 78.0 | 45 | 54.9 | 28 | 34.2 | 24 | 29.3 |
|  | 5-14 | 163 | 125 | 76.7 | 83 | 50.9 | 65 | 39.9 | 40 | 24.5 |
|  | 15-39 | 141 | 93 | 66.0 | 51 | 36.2 | 42 | 29.8 | 29 | 20.6 |
|  | ≥40 | 42 | 22 | 52.4 | 6 | 14.3 | 12 | 28.6 | 10 | 23.8 |
| **Relation to the study infant** | Self | 47 | 37 | 78.7 | 27 | 57.5 | 13 | 27.7 | 15 | 31.9 |
|  | Sibling | 162 | 124 | 76.5 | 87 | 53.7 | 55 | 34.0 | 40 | 24.7 |
|  | Cousin | 124 | 91 | 73.4 | 56 | 45.2 | 51 | 41.1 | 28 | 22.6 |
|  | Mother | 46 | 27 | 58.7 | 16 | 34.8 | 13 | 28.6 | 11 | 23.9 |
|  | Father | 30 | 17 | 56.7 | 7 | 23.3 | 7 | 23.3 | 5 | 16.7 |
|  | Other HH members | 74 | 50 | 67.6 | 22 | 29.7 | 24 | 32.4 | 20 | 27.0 |
| **Sex** | Female | 269 | 186 | 69.1 | 121 | 45.0 | 89 | 33.1 | 65 | 24.2 |
|  | Male | 214 | 160 | 74.8 | 94 | 43.9 | 74 | 34.6 | 54 | 25.2 |
| **School going** | No | 314 | 217 | 69.1 | 128 | 40.8 | 104 | 33.1 | 76 | 24.2 |
|  | Yes | 169 | 129 | 76.3 | 87 | 51.5 | 59 | 34.9 | 43 | 25.4 |
| **Number of individuals per HH (household sizes)** | 4 to 7 | 95 | 75 | 79.0 | 50 | 52.6 | 37 | 39.0 | 21 | 22.1 |
|  | 8 to10 | 120 | 77 | 64.2 | 53 | 44.2 | 29 | 24.2 | 26 | 21.7 |
|  | 11 to 16 | 144 | 97 | 67.4 | 48 | 33.3 | 49 | 34.0 | 40 | 27.8 |
|  | 17 to 37 | 124 | 97 | 78.2 | 64 | 51.6 | 48 | 38.7 | 32 | 25.8 |

Symptomatic individuals contributed to 410 (32.2%) of the 1274 samples that were positive for any HCoV and 3150 (20.1%) of 15645 hCoV negative samples. Symptomatic individuals contributed to 240 (36.9%), 132 (31.6%) and 47 (19.5%) of the total number of samples that tested positive for OC43, NL63 and 229E, respectively, and correspondingly 3320 (20.4%), 3428 (20.8%) and 3513 (21.1%) of samples that tested negative. There was a statistically significant association between the presence of respiratory symptoms during sampling times and detection of any hCoV (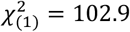, p-value <0.001), OC43 (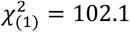, p-value <0.001) and NL63 (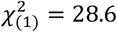, p-value <0.001) but not 229E (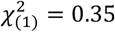, p-value =0.555).

### HCoV infection episodes

The pattern of shedding of each of the three hCoV types and of all hCoVs, is displayed in Figure 1. Over the study period, 346 (71.6%) of the 483 individuals experienced one or more hCoV infection episode. The total number of individual infection episodes was 260 for OC43, 216 for NL63, 140 for 229E and 629 for any hCoV type. Of these the number of episodes symptomatic was 116 (44.5%) for OC43, 85 (39.4%) for NL63, 35 (25.0%) for 229E and 228 (36.3%) for any hCoV type. The proportion of symptomatic episodes differed by hCoV type (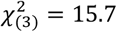, p-value =0.001) and by age for any hCoV type (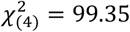, p-value < 0.001) and each of the three hCoV types (OC43; 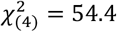, p-value <0.001, 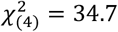, p-value <0.001 and 229E; (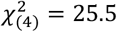, p-value <0.001). Of the total episodes, 29 (11.2%), 51 (23.6%), 28 (20%) and 105 (16.7%) of OC43, NL63, 229E or any hCoV infection episodes, respectively were either right or left-censored and were excluded in survival analysis.

**Figure 1:**
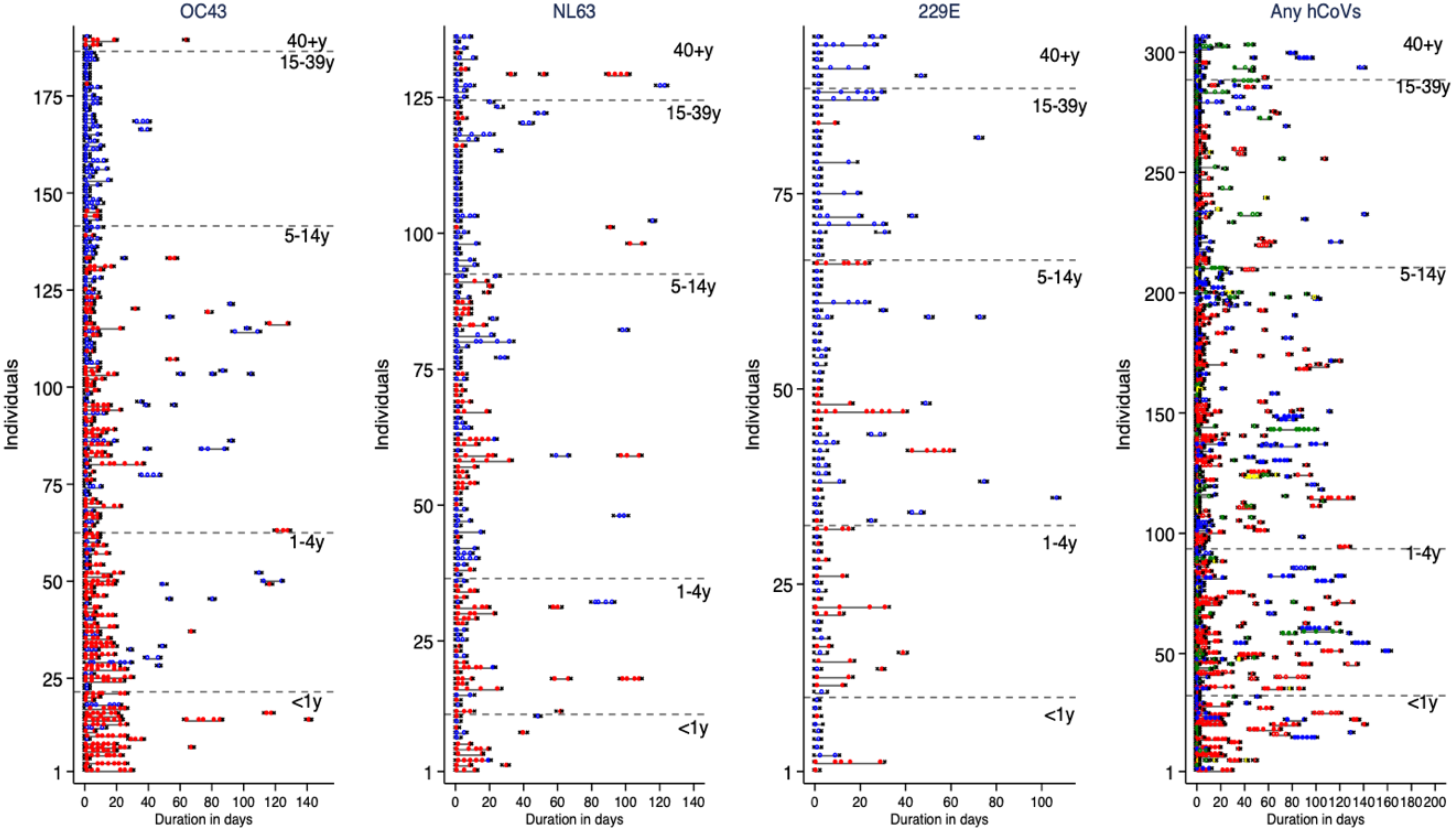
Individual infection episodes showing the duration of shedding hCoV type. *The grey lines indicate duration of the episodes, dots indicate positive samples within an episode and the crosses at both ends of the line denote the start and end of the infection episodes. For the first three graphs (OC43, NL63 and 229E) symptomatic episodes are shown by red filled dots while asymptomatic episodes are shown by blue unfilled dots. In the final graph, red, blue, green and yellow dots indicate samples positive for OC43, NL63, 229E and HCoV-HCoV coinfection. Filled dots indicate symptomatic episodes while unfilled dots are asymptomatic episodes*.

On average, the peak viral load of the individual infection episodes was higher in symptomatic compared to asymptomatic episodes (Figure 2). This was supported by linear regression model, adjusted for age, fitted on the peak viral load showing symptomatic infection episodes had a higher viral load (lower Ct values) compared to asymptomatic episodes for OC43 *(β* = −4.16, 95% CI= -5.55, -2.77, p-value<0.001), NL63 (*β* = −2.74, 95% CI= -5.06, -0.43, p-value=0.020), 229E (*β* = −5.79, 95% CI= -9.88, -1.70, p-value=0.006) and any hCoV (*β* = −3.11, 95% CI= -4.50, -1.72, p-value <0.001).

**Figure 2:**
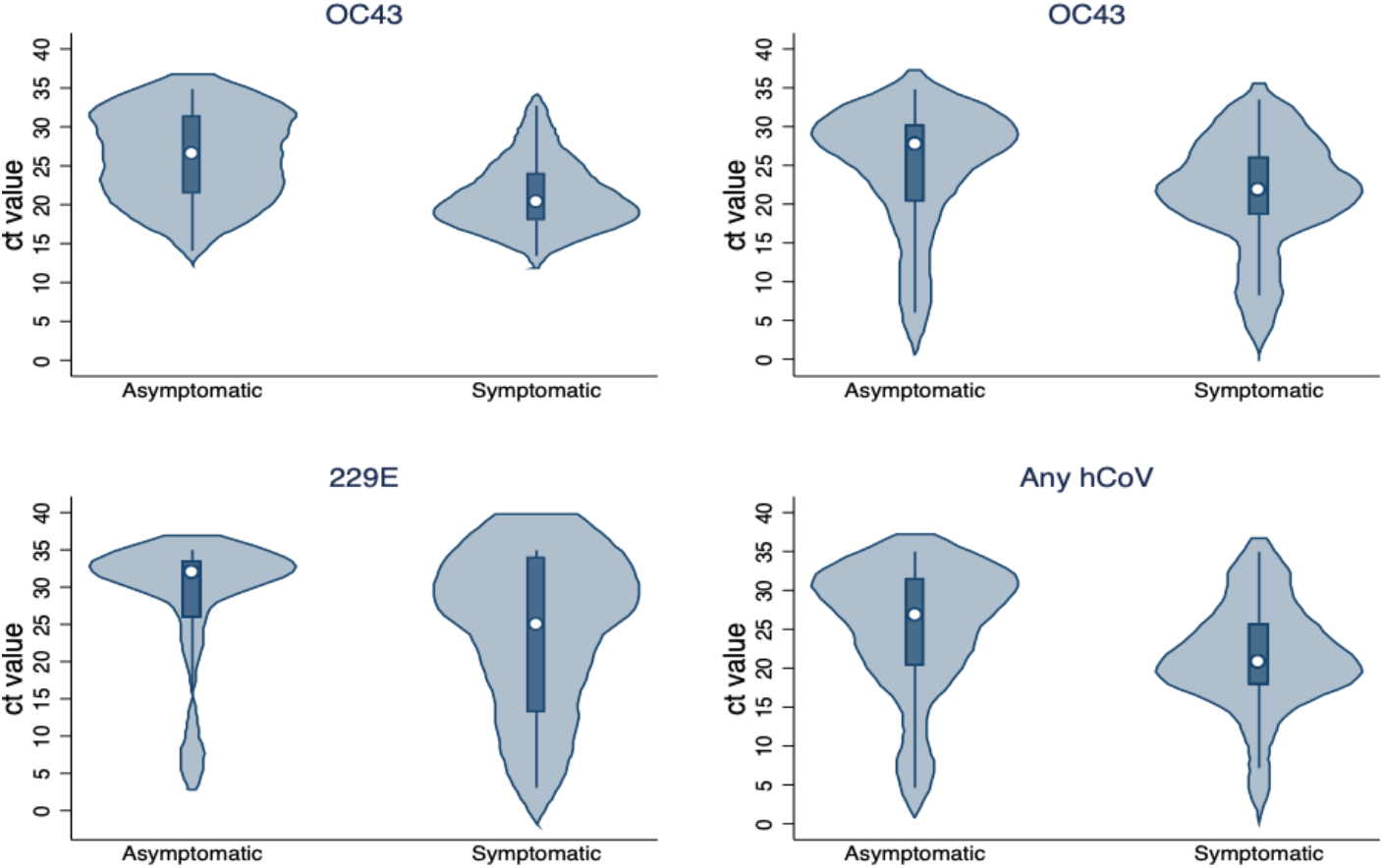
Violin plots showing the distribution of the peak viral load (expressed in Ct values) across infection episodes by hCoV type and symptomatic status.

### Durations of hCoV shedding and factors influencing shedding patterns

The duration of virus shedding varied by hCoV type with the longest median duration observed for OC43 (7.5 days, IQR: 3.5 -13.5) and the shortest median duration for 229E (3.5 days, IQR: 3.5 - 7.0) (Figure 3). The results of the survival analysis of shedding durations are shown in Table 2. For OC43, the rate of recovery from an infection episode, compared to children aged <1 year, was higher in children aged 5-14 years (aHR=2.21, 95% CI: 1.40 – 3.47, p-value=<0.001), those aged 15-39 years (aHR=1.72, 95% CI: 1.05 – 2.83, p-value=0.032) and older adults 40 years or more (aHR=3.44, 95% CI: 1.97 – 6.01, p-value <0.001). Age dependence for duration of infection episodes was not observed for NL63, 229E and pooled hCoV type analysis. Lower rates of recovery from OC43 virus shedding (i.e. longer shedding durations) were observed among symptomatic (compared to asymptomatic) individuals (aHR=0.56, 95% CI: 0.39 - 0.81, p-value=0.002), and individuals with medium peak viral load (aHR=0.32, 95% CI: 0.22-0.47, p-value <0.001) and high peak viral load (aHR= 0.22, 95% CI: 0.14-0.36, p-value <0.001) compared to individuals with low peak viral load. Recovery rates from NL63 infection episode was dependent on detection of other viral pathogens within the same infection episode (aHR= 0.54, 95% CI: 0.34 - 0.87, p-value= 0.011), being symptomatic (aHR= 0.67, 95% CI: 0.50 - 0.90, p-value=0.009), and medium (aHR= 0.49, 95% CI: 0.33 - 0.74, p-value=0.001) and high (aHR=0.37, 95% CI: 0.24 - 0.59, p-value <0.001) peak viral load in an infection episode. For 229E, having medium (aHR=0.49, 95% CI: 0.30 - 0.78, p-value=0.003) and high (aHR=0.21, 95% CI: 0.11 - 0.40, p-value=<0.001) peak viral load significantly affected the rate of recovery from an infection episode. The KM curves for each factor are presented in supplementary figures S2-S5.

**Table 2:** Factors influencing the recovery rate of hCoV infections from multivariable cox proportional hazards model analysis.

| Characteristics | Categories | OC43 |  | NL63 |  | 229E |  | Any hCoV |  |
| --- | --- | --- | --- | --- | --- | --- | --- | --- | --- |
|  |  | aHR<br>(95% CI) | p-value | aHR<br>(95% CI) | p-value | aHR<br>(95% CI) | p-value | aHR<br>(95% CI) | p-value |
| Sex | Female | Ref |  | Ref |  | Ref |  | Ref |  |
|  | Male | 1.06<br>(0.83 - 1.34) | 0.634 | 1.22<br>(0.90 - 1.66) | 0.201 | 0.90<br>(0.64 - 1.26) | 0.549 | 1.07<br>(0.91 - 1.27) | 0.397 |
| Age group (in years) | <1 y | Ref |  | Ref |  | Ref |  | Ref |  |
|  | 1-4 y | 1.49<br>(0.96 - 2.30) | 0.075 | 0.60<br>(0.28 - 1.28) | 0.186 | 1.00<br>(0.47 - 2.13) | 0.999 | 1.08<br>(0.75 - 1.56) | 0.672 |
|  | 5-14 y | 2.21<br>(1.40 - 3.47) | <0.001 | 0.69<br>(0.32 - 1.48) | 0.341 | 1.04<br>(0.57 - 1.95) | 0.877 | 1.31<br>(0.92 - 1.88) | 0.143 |
|  | 15-39 y | 1.72<br>(1.05- 2.83) | 0.032 | 0.99<br>(0.46 - 2.12) | 0.976 | 0.79<br>(0.39 - 1.58) | 0.504 | 1.13<br>(0.76 - 1.68) | 0.552 |
|  | ≥40 y | 3.44<br>(1.97 – 6.01) | <0.001 | 0.95<br>(0.43 – 2.09) | 0.902 | 0.90<br>(0.47 – 1.71) | 0.739 | 1.27<br>(0.76 – 2.11) | 0.370 |
| Presence of other respiratory viruses | No | Ref |  | Ref |  | Ref |  | Ref |  |
|  | Yes | 0.98<br>(0.73 - 1.32) | 0.902 | 0.54<br>(0.34 - 0.87) | 0.011 | 0.70<br>(0.47 - 1.04) | 0.077 | 0.80<br>(0.65 - 0.97) | 0.026 |
| **Symptomatic** | No | Ref |  | Ref |  | Ref |  | Ref |  |
|  | Yes | 0.56<br>(0.39 - 0.81) | 0.002 | 0.67<br>(0.50 - 0.91) | 0.009 | 0.71<br>(0.47 - 1.07) | 0.103 | 0.62<br>(0.51 - 0.76) | <0.001 |
| **Peak viral load** | Low | Ref |  | Ref |  | Ref |  | Ref |  |
|  | Medium | 0.32<br>(0.22-0.47) | <0.001 | 0.49<br>(0.33 - 0.74) | 0.001 | 0.49<br>(0.30 - 0.78) | 0.003 | 0.44<br>(0.34 - 0.57) | <0.001 |
|  | High | 0.22<br>(0.14-0.36) | <0.001 | 0.37<br>(0.24 - 0.59) | <0.001 | 0.21<br>(0.11 - 0.40) | <0.001 | 0.31<br>(0.23 - 0.42) | <0.001 |
| **HCoV type** | OC43 | – | – | – | – | – | – | Ref |  |
|  | NL63 | – | – | – | – | – | – | 1.17<br>(0.95 – 1.43) | 0.125 |
|  | 229E | – | – | – | – | – | – | 1.02<br>(0.78 – 1.35) | 0.861 |
|  | hCoV – hCoV<br>coinfection | – | – | – | – | – | – | 2.64<br>(1.97 – 3.55) | <0.001 |

Key: aHR, adjusted Hazard Ratio; hCoV-hCoV coinfection denotes infection episodes in which an individual tested positive for two or more hCoVs.

**Figure 3:**
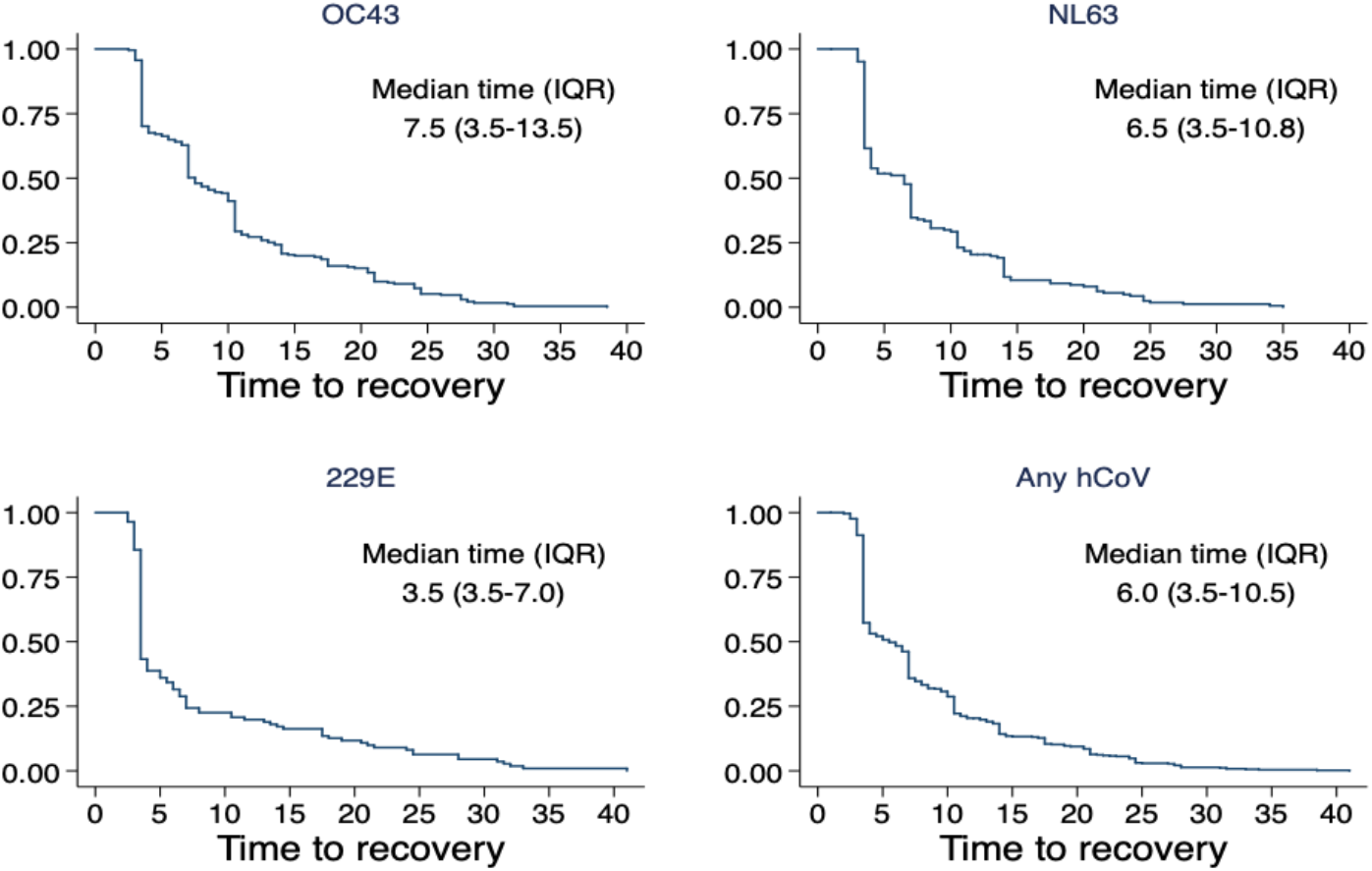
Time to recovery of hCoV infections for OC43, NL63, NL63 and any hCoV based on survival analysis (Kaplan Meir curves)

### Duration of symptomatic period

The median duration of symptoms (Supplementary Table S1) was 7.0, 4.0, 3.5 and 4.0 days, for OC43, NL63, 229E and any hCoV, respectively. For any hCoV median durations tended to decline with increasing age, increase in the presence of other respiratory viruses and increase for high peak viral load. There was some variation between hCoV type.

The rate of recovery from symptoms (Supplementary Table S2) within any hCoV infection episodes increased with age; 1-4 years (aHR=1.45, 95% CI: 1.03 - 2.04, p-value=0.034), 5-14 years (aHR=1.64, 95% CI:1.12 - 2.39, p-value=0.010), 15-39 years (aHR=2.25, 95% CI: 1.30 – 3.89, p-value=0.003); 40 years and above (aHR=1.61, 95% CI: 1.10 – 2.37, p-value=0.014). A similar pattern was observed on OC43 alone out of the three hCoVs. Infection episodes with a coinfection with another virus had lower rates of recovery from symptoms for 229E (aHR=0.33, 95% CI: 0.17 - 0.63, p-value=0.001) and any hCoV (aHR=0.71, 95% CI: 0.55 - 0.91, p-value=0.006). The rate of symptoms clearance within OC43 infection episodes was influenced by medium peak viral load (aHR=0.15, 95% CI 0.07 - 0.31, p-value < 0.001) and high peak viral load (aHR=0.14 95% CI: 0.07 - 0.29, p-value<0.001). Similarly, a lower recovery rate was observed for any hCoV infection episodes with medium peak viral load (aHR 0.44, 95% CI: 0.29 - 0.68, p-value < 0.001) and high peak viral load (aHR=0.35, 0.23 - 0.54, p-value<0.001).

### Reinfection

Details of reinfections in the study cohort are given in Table 3. Of 483 study individuals, 346 (71.6%) experienced one or more infection episodes and 171 (35%) experienced multiple infection episodes for any hCoV type. Of the 215, 163, and 119 individuals who had first infections of OC43, NL63 and 229E, respectively, a corresponding 35 (16.3%), 44 (27.0%) and 20 (16.8%), experienced one or more homologous reinfections. In summation, a total of 497 first infections with OC43, NL63 and 229E were observed of which 99 (19.9%) were infected at least once with the homologous type. Of the 629 episodes, 346 (55.0%) were first and 283 (45.0%) secondary, infections and 154/283 (54.4%) reinfections were of homologous type. Analysing the total infections by type, the proportion of homologous reinfection episodes varied 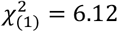, p-value =0.047) by type (17.3% of 260 OC43 episodes, 24.5% of 216 NL63 episodes and 15.0% of 140 episodes due to 229E). In addition, 48 (31.2%) of the 154 homologous reinfection episodes due to any hCoV were symptomatic, compared to 56 (43,4%) of the 129 heterologous reinfections. There was no difference 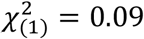, p-value =0.764) in the median time to reinfection between homologous (41 days, IQR: 19-73 days) and heterologous (40 days, IQR: 23-60 days) infection episodes. In the period between end of the first infection episode and start of the first reinfection episode, a total of 16 (9.4%) did not have any swabs taken, possibly indicating a continuation of the first episode, while 9 (5.3%), 8 (4.7%), 14 (8.2%) and 124 (72.5%) had 1,2,3 and ≥4 PCR negative samples respectively out of the 171 individuals with multiple infections.

**Table 3:** **Frequency distribution of individual infection episodes by hCoV type in 483 individuals from a household cohort in rural Kenya.**

|  | Type | 1st | 2nd | 3rd | 4th | 5th | 6<sup>th</sup> | 7th | Total reinfection | Total infection |
| --- | --- | --- | --- | --- | --- | --- | --- | --- | --- | --- |
| **Individual hCoV types episodes** | OC43 | 215 | 35 | 9 | 1 | — | — | — | 45 | 260 |
|  | NL63 | 163 | 44 | 8 | 1 | — | — | — | 53 | 216 |
|  | 229E | 119 | 20 | 1 | — | — | — | — | 21 | 140 |
|  | All types summation | 497 | 99 | 18 | 2 | — | — | — |  |  |
| **Any hCoV type episodes** | Any Homologous |  | 66 | 52 | 25 | 8 | 2 | 1 | 154 |  |
|  | Any infection | 346 | 171 | 73 | 27 | 9 | 2 | 1 | 283 | 629 |

There was no difference in the proportion of symptomatic episodes in the first infection episodes (124/346, 35.8%) compared to reinfections (104/283, 36.8%) (p-value=0.7952). Disaggregation of symptomatic infection episodes by hCoV type showed that the contribution of each pathogen to the first 124 symptomatic episodes varied; OC43 (58.1%), NL63 (26.6%), 229E (14.5%) and hCoV-hCoV coinfection (0.8%). Similarly, out of the 104 symptomatic reinfections episodes, there was a variation in the proportion of hCoV type; OC43 (36.5%), NL63 (44.2%), 229E (12.5%) and hCoV-hCoV coinfection (6.7%).

### Transmission of hCoV in households

All the 47 households had at least one of the three hCoV detected while hCoV-OC43, NL63 and 229E were detected in 44 (93.6%), 33 (70.2%) and 30 (63.8%) of the households, respectively. There were 78, 48, 59 and 201 household introductions for OC43, NL63, 229E and any hCoV, respectively. Siblings and cousins, predominantly of school going age, to the study infants were index cases for 59.0%, 64.6%, 52.5% and 53.7% of OC43, NL63, 229E and any hCoV household introductions, respectively (Figure 4). Out of the total number of household introductions, 46 (59.0%), 30 (62.5%), 23 (39.0%) and 96 (47.8%) led to secondary infections in 35, 23, 17 and 43 households for OC43, NL63, 229E and any hCoV, respectively. The proportion of secondary infections due to the 3 hCoV types was significantly different across types (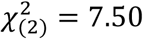, p-value =0.023).

**Figure 4:**
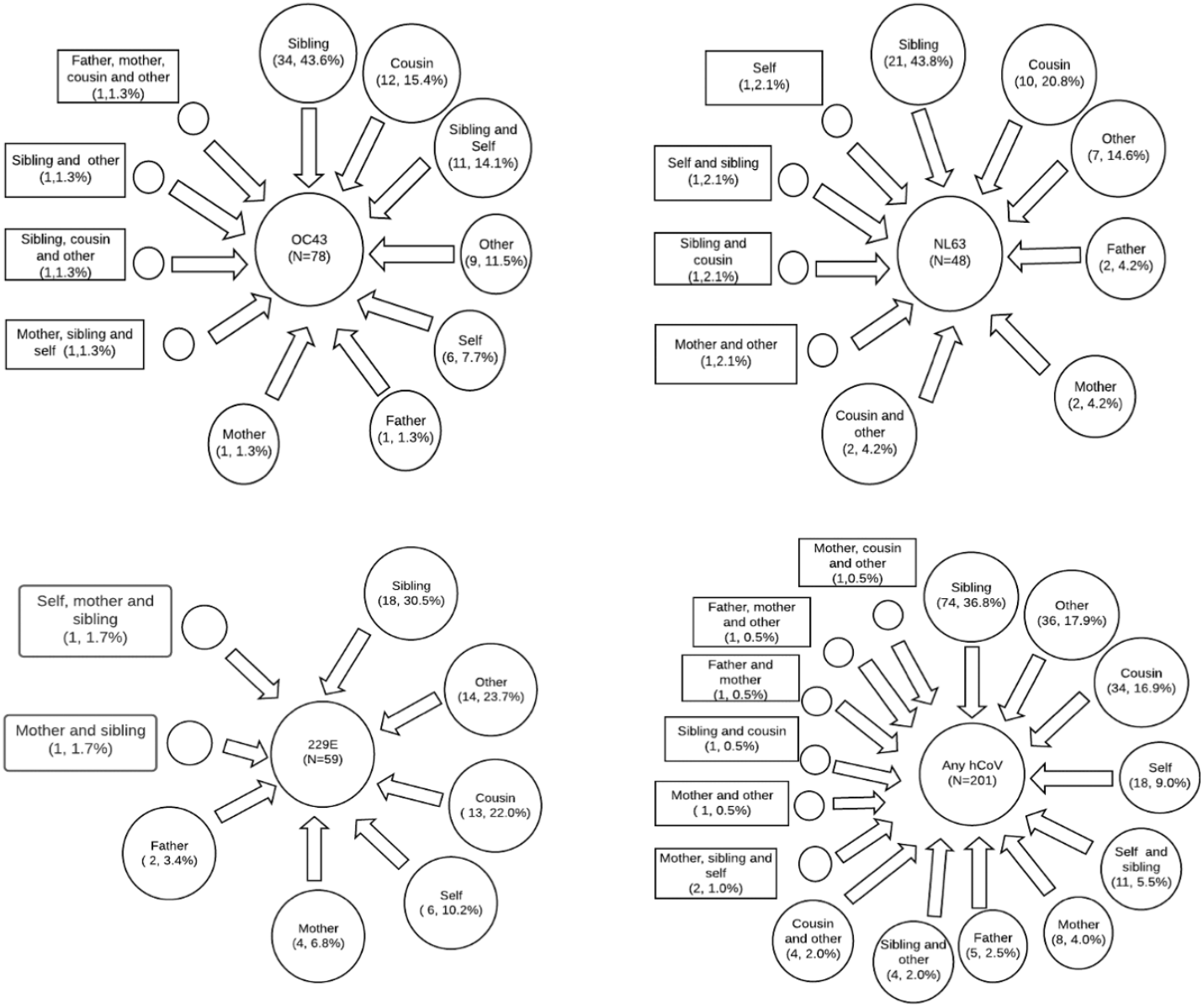
Frequency distribution of who brings hCoV infections in households by HCoV type.

The risk of generating a secondary case after introduction of any of the three endemic hCoV in the household was higher for index cases whose peak viral load was medium (aOR=5.29, 95% CI: 2.34 – 11.96, p-value <0.001) or high (aOR=8.12, 95% CI: 2.92 - 22.51, p-value <0.001) compared those with a low peak viral load. However, being a symptomatic index case was not associated with increased risk of infecting other members of the household (aOR=0.97, 95% CI: 0.42 - 2.21, p-value=0.933) compared to asymptomatic index cases (Supplementary Table S3).

## Discussion

Longitudinal studies of households have played an important role in developing understanding of the epidemiology of respiratory viruses [7,14]. Here we continue this approach, reporting an intensive surveillance of 483 household members in rural coastal Kenya [11], to delineate the natural history of infection and transmission patterns of three endemic coronaviruses (OC43, NL63 and 229E). This involved the application of sensitive molecular diagnostic methods [7,14], and additionally applied sampling that was frequent and irrespective of observed symptoms [8]. The hCoV types were common in this setting with each of the 47 households, and about 72% of the enrolled household members, experiencing infection with at least one of three targets over the six months of the study. A note of caution in interpreting the results of this study is that infection status determined by PCR assay is not necessarily indicative of active infection or an individual’s infectiousness.

Crude attack rates were highest for hCoV-OC43 and lowest for 229E, higher in general for younger age classes (<15 years of age), school-age children and for males. These results are broadly consistent with the findings by Monto *et al*. who also found highest incidence for OC43, lowest for 229E, and higher incidence among those aged below 5 years for NL63 and OC43 [7].

The three hCoV types had differing durations of shedding ranging from 3.5 days (229E) to 7.5 days (OC43). However, these median time estimates are influenced by our sampling frequency: predominantly every 3-4 days. The duration of shedding was longer in episodes with high peak virus load and which were symptomatic. Consistent with findings from other studies, we report occurrence of hCoV infection episodes among asymptomatic individuals [15–17] who had lower viral load [18] and shorter durations of virus shedding compared to symptomatic episodes. Despite asymptomatic infections being predominant (>70% of episodes) the above findings suggest they were less likely to transmit infection compared to symptomatic individuals. The duration of symptomatic episodes was related to peak virus load as reported elsewhere [25] and tended to decline with increasing age.

Participants of all ages had appreciable risk of infection for the three endemic viruses suggesting previous infection does not provide solid immunity. This is supported by our observation that, within the short period of the study, reinfections were common and as frequently of homologous as heterologous type. Overall, 20% of individuals with a first infection of one or other type, were reinfected by the same type at least once, most commonly for type NL63 (24.5%). Homologous reinfections were frequently (>30%) symptomatic. We report no difference in the proportion of symptomatic cases between the first episodes and reinfection episodes and note that the time to reinfection with homologous was similar to heterologous episodes (∼40 days). Our observations indicate that immunity to reinfection is commonly short lived and does not appear to be type specific. A recent serological study involving 10 adult men detected reinfections from seasonal coronaviruses but most frequently occurring after an interval of 12 months[19]. A limitation of our analysis is that reinfections might in fact have been prolonged shedding from a single infection. This is likely not a major effect as in most presumed reinfections (>70%) there were at least 4 PCR test negative results between episodes.

Older children (siblings and cousins) and other adults were the major introducers of hCoV transmission into the household compared to RSV transmission in the same households whereby older children (> 32%) were the leading primary cases [11]. Similarly, children have been reported to form the highest proportion of index cases in the USA and UK [7,20]. However, presence of older adults, children, smokers and individuals with chronic ailments within the households in the UK study was associated with increased household transmission [20]. Secondary transmission of hCoV to other household members upon introduction was high (48%) for any of the three hCoVs (ranged from 39% to 62% across type). This differs from a recent study in England which concluded that the vast majority (>90%) of observed hCoV infections were acquired outside the household [20]. In our study, the risk of secondary transmission was higher among index cases with high viral loads. Interestingly, there was no significant association between the presence of symptoms among index cases and the risk of secondary transmission, as observed elsewhere [20].

In conclusion, endemic coronaviruses are common within the household setting, infecting all age groups, and often without eliciting symptoms. Secondary transmission following household introduction is associated with viral load but not, it appears, with symptomatic status, and homologous reinfection is common for all hCoV types.

## Data Availability

The replication data and analysis scripts for this manuscript has been made available at the Harvard Dataverse: (https://doi.org/10.7910/DVN/CPJ9B4). Some of the clinical dataset contains potentially identifying information on participants and is stored under restricted access. Requests for access to the restricted dataset should be made to the Data Governance Committee of the KEMRI-Wellcome Trust Research Programme.

## Supplementary figures legend

**Supplementary Figure S1:**
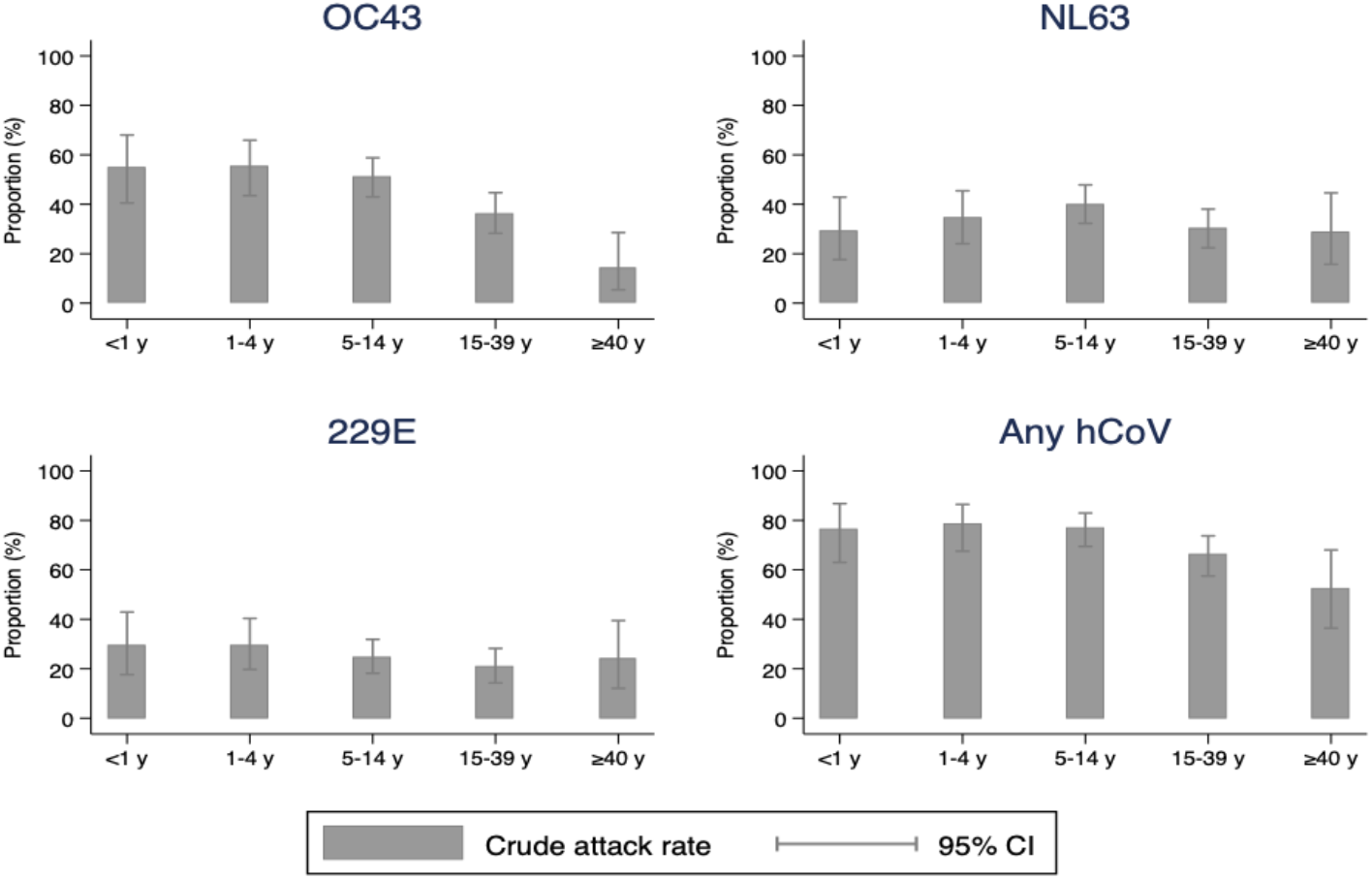
Crude attack rates by age category for each hCoV.

**Supplementary Figure S2:**
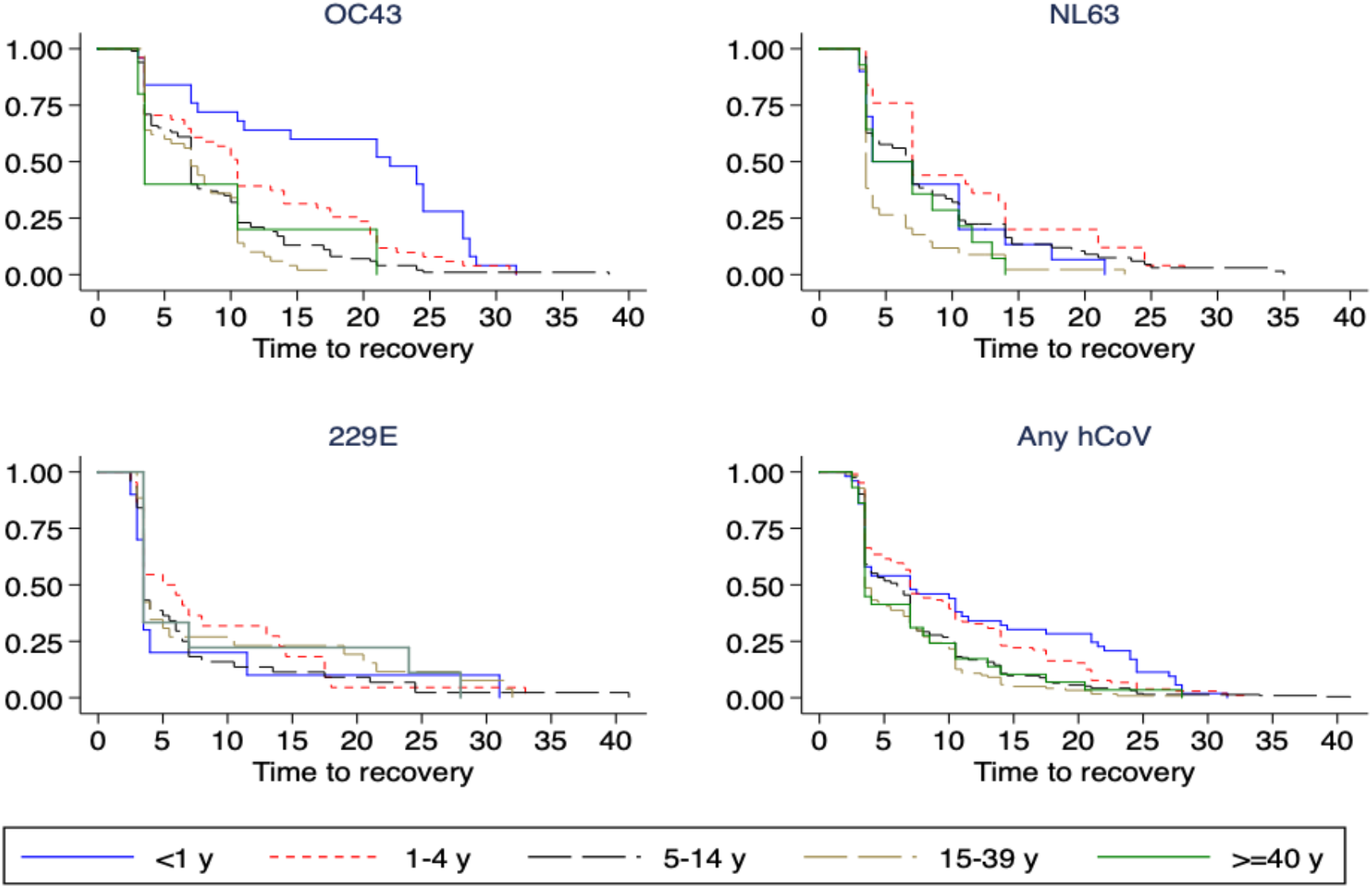
Kaplan Meir curves on virus shedding by age category across hCoV type.

**Supplementary Figure S3:**
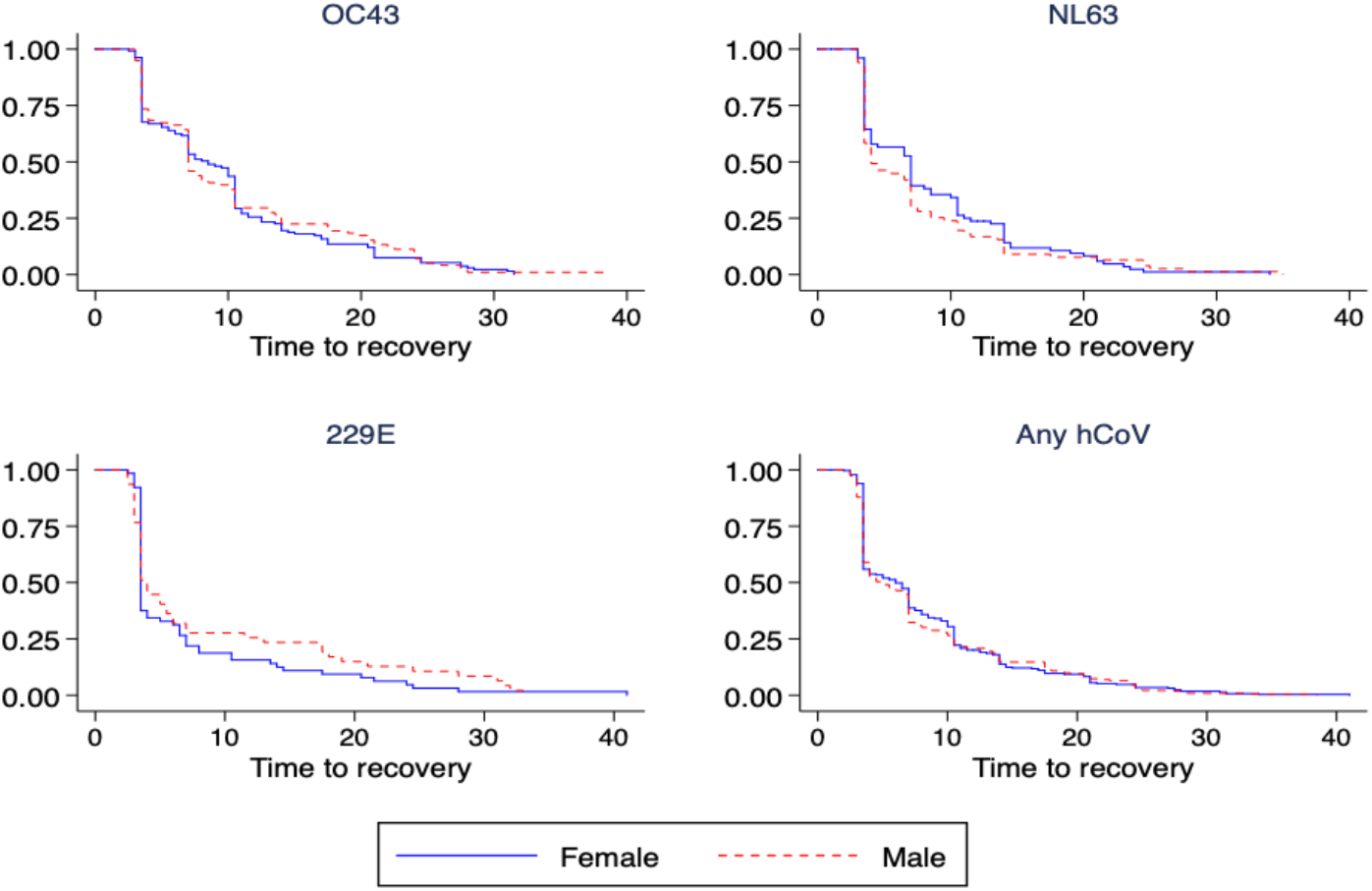
Kaplan Meir curves on virus shedding by sex category across hCoV type.

**Supplementary Figure S4:**
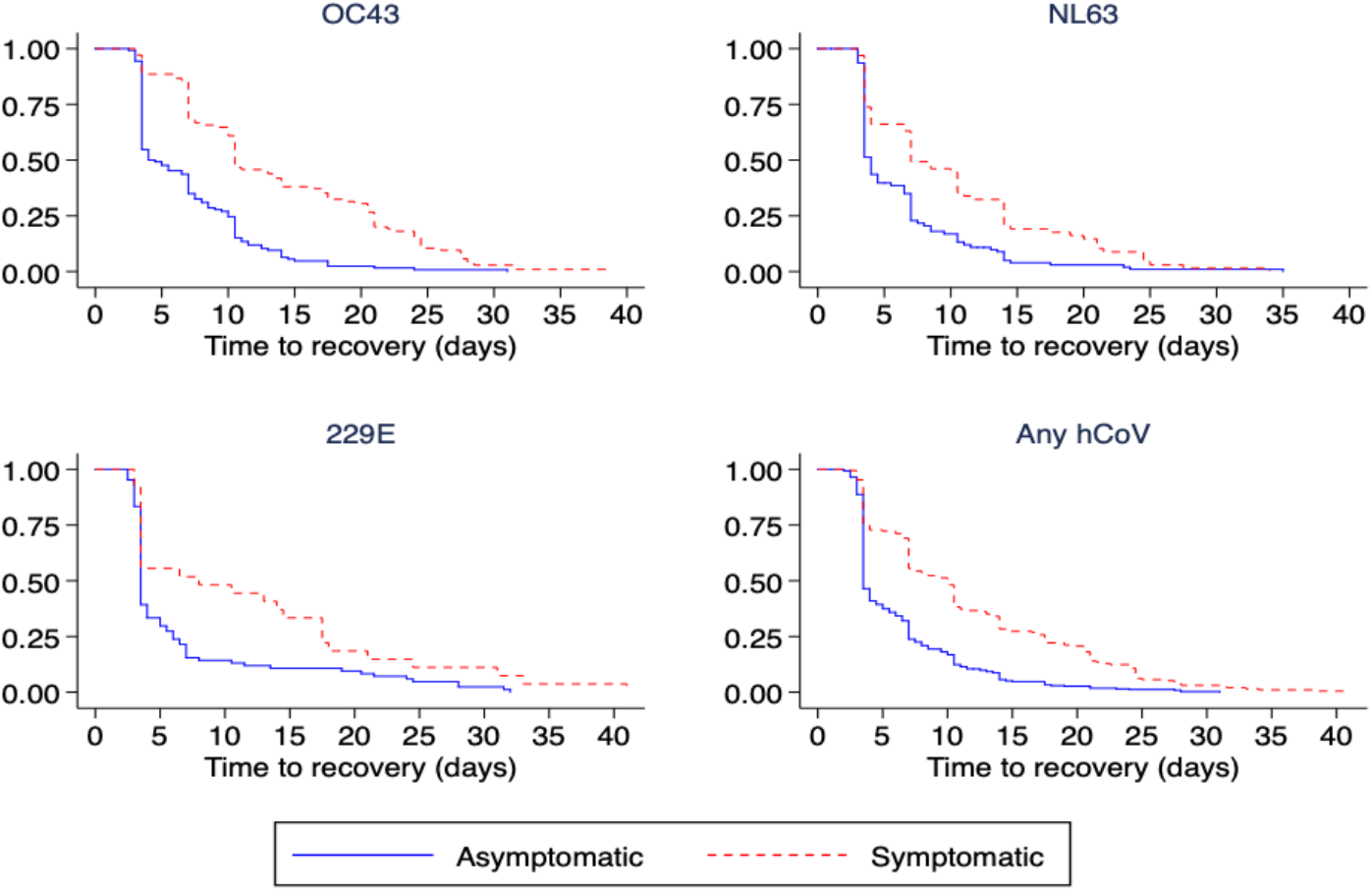
Kaplan Meir curves on virus shedding by symptomatic status category across hCoV type.

**Supplementary Figure S5:**
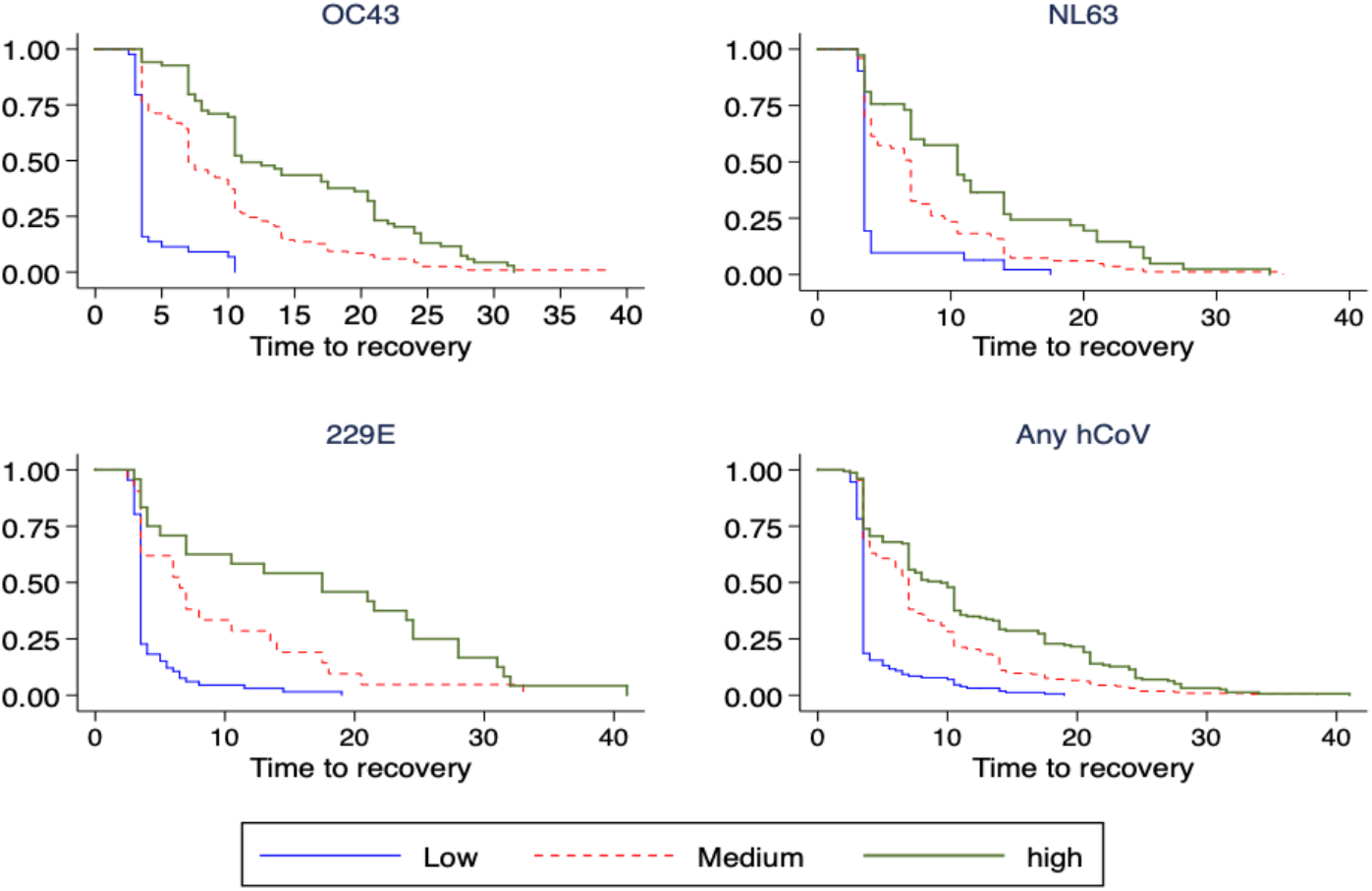
Kaplan Meir curves on virus shedding by category of peak viral load across hCoV type.

